# Beta Burst Waveform Motifs Link Cortico-Subcortical Connectivity and Clinical Improvement in Parkinson’s Disease

**DOI:** 10.1101/2025.07.09.25331208

**Authors:** Hasnae Agouram, Maciej J Szul, Alexandre Eusebio, Emmanuel Daucé, Pierpaolo Sorrentino, James J Bonaiuto

## Abstract

Beta bursts are increasingly being studied as biomarkers of Parkinson’s disease (PD) symptoms and therapeutic response. While most prior work has focused on time-frequency burst features such as duration or amplitude, recent evidence suggests that the temporal waveform of individual bursts carries additional functional significance. Here, we investigated whether distinct beta burst waveform motifs in the subthalamic nucleus (STN) and sensorimotor cortex are modulated by levodopa and whether these modulations relate to cortico-subcortical connectivity and clinical improvement. We analyzed resting-state activity from 11 PD patients recorded from deep contacts in the STN and EEG electrodes over the motor cortex, both before and after levodopa administration. Using a novel adaptive burst detection algorithm, we extracted individual beta bursts and characterized their waveform variability using principal component analysis. While levodopa had inconsistent effects on conventional time-frequency burst features, it selectively modulated the rate of specific burst waveform motifs in the sensorimotor cortex, consistently across patients. Furthermore, we found that levodopa increased functional connectivity between the STN and cortex specifically during bursts with certain waveform motifs, and these connectivity changes predicted individual clinical improvement. Our findings suggest that beta bursts are not uniform events but comprise a set of functionally distinct motifs whose rates and network involvement are modulated by dopaminergic therapy. This waveform-based approach refines our understanding of pathological beta activity in PD and offers novel targets for personalized, adaptive neuromodulation strategies.

## Introduction

Parkinson’s disease (PD) is a neurodegenerative disorder characterized by bradykinesia, muscle rigidity, and resting tremor, resulting from dysfunction in basal ganglia-cortical circuits (Jankovic, 2008; Poewe et al., 2017). Aberrant beta-band activity (13-35 Hz) in the basal ganglia is a hallmark of PD, with excessive beta power strongly linked to motor impairment (Brown, 2003; Kühn et al., 2006; Chen et al., 2010; Eusebio et al., 2011; Özkurt et al., 2011; Neumann et al., 2016; Oswal et al., 2016; Trager et al., 2016). Suppression of pathological beta activity in the subthalamic nucleus (STN), whether pharmacologically with levodopa or via deep brain stimulation (DBS), has been shown to correlate with clinical improvement (Kühn et al., 2006, 2008, 2009; Chen et al., 2010; López-Azcárate et al., 2010; Eusebio et al., 2011; Özkurt et al., 2011; Little et al., 2016; Oswal et al., 2016; Trager et al., 2016). However, sensorimotor beta activity occurs mainly in transient bursts rather than continuous oscillations (Murthy and Fetz, 1992, 1996; Kilner et al., 2003; Feingold et al., 2015), and the duration of these bursts has emerged as a stronger predictor of clinical impairment than average beta power. This has motivated the development of closed-loop DBS systems that deliver stimulation contingent on the presence of prolonged beta bursts (Tinkhauser et al., 2017b, 2017a; Anidi et al., 2018; Kehnemouyi et al., 2021). However, such strategies typically rely on time-frequency features that account for only a limited fraction of symptom variability (Lofredi et al., 2023), suggesting that clinically-relevant aspects of beta activity, potentially those related to burst waveform structure, remain unexplored in current models of PD pathophysiology and neuromodulation.

In the sensorimotor cortex, the shape of individual beta bursts in the temporal domain exhibits considerable variability, which may index functionally distinct neural processes (Howe et al., 2011; Sherman et al., 2016; Karvat et al., 2020; Kosciessa et al., 2020; Rayson et al., 2023; Szul et al., 2023). Recent work has shown that beta bursts with different waveform shapes display distinct rate dynamics during movement, suggesting that they arise from separate underlying neural mechanisms (Rayson et al., 2023; Szul et al., 2023). However, most prior studies have treated beta bursts as uniform, focusing on their rate, timing, or average waveform shape, while largely overlooking the diversity in their spectral and temporal features (though see Zich et al., 2020; Duchet et al., 2021; Enz et al., 2021). These findings raise the possibility that sensorimotor beta activity comprises a repertoire of burst types, each with distinct dynamical properties and potential computational roles, offering a new lens through which to reconcile competing accounts of beta’s function in motor control.

Because beta bursts dynamics are shaped by recurrent interactions within cortico-basal ganglia loops (Tinkhauser et al., 2018; Cagnan et al., 2019; Yu et al., 2021; Yao et al., 2025), diversity in beta burst waveform morphology is unlikely to be confined to the cortex alone. It is therefore plausible that similar diversity exists in the basal ganglia, particularly the STN. Systematically characterizing burst waveform motifs across both cortical and subcortical areas may yield new insights into the network-level mechanisms of PD and identify more specific biomarkers of dopaminergic state and clinical response.

Previous studies investigating network-level functional connectivity (FC) within the basal ganglia motor circuit (BGMC) in PD patients have consistently shown that dopaminergic depletion impairs cortico-subcortical interactions (Haslinger et al., 2001; Wu and Hallett, 2005; Herz et al., 2014; Gao et al., 2016; Sorrentino et al., 2021) and that levodopa partially restores these network dynamics (Haslinger et al., 2001; Buhmann et al., 2003; Kraft et al., 2009; Palmer et al., 2009; Wu et al., 2009; Martinu et al., 2012). However, most of these investigations have relied on indirect or temporally coarse measures such as fMRI (Bowyer, 2016), or on model-free connectivity metrics that are poorly suited for capturing the brief, event-related nature of beta bursts. Because beta bursts are discrete and transient events, with variable waveform shapes that may reflect functionally distinct network states, a time-resolved, burst-specific approach is required to elucidate how levodopa alters communication within the BGMC. In particular, it remains unknown whether the reorganisation of cortico-subcortical connectivity by dopaminergic medication is selective for bursts with particular waveform motifs, a possibility that would support a more granular, physiologically grounded framework for understanding levodopa’s therapeutic effects.

In this study, we analyzed simultaneously recorded STN-LFP and scalp EEG activity from 11 PD patients implanted with DBS electrodes, both OFF and ON levodopa. Using a recently developed adaptive burst detection algorithm (Rayson et al., 2023; Szul et al., 2023), we extracted individual beta bursts from both sensorimotor cortex and STN, and characterized their waveform shapes via principal component analysis (PCA). We found that while conventional time-frequency features showed inconsistent modulation by medication, levodopa selectively altered the rate of distinct burst waveforms motifs in the sensorimotor cortex. These waveform-specific modulations were accompanied by selective increases in Granger causality between the STN and sensorimotor cortex, specifically during bursts within certain motifs, and this connectivity change predicted levodopa-induced individual clinical improvement. Our results reveal that beta burst waveform motifs serve as dynamic indices of functional brain state and dopaminergic response, offering a novel, mechanistically grounded metric for monitoring disease progression and informing adaptive neuromodulation strategies.

## Results

### Time-frequency burst features are not consistently altered by levodopa

We used a recently developed adaptive, iterative algorithm to detect all beta burst events across a broad range of amplitudes, without relying on fixed thresholds (Rayson et al., 2023; Szul et al., 2023). Bursts were extracted from the C3 and C4 EEG electrodes over the sensorimotor cortex, as well as from the STN contacts with the highest beta power in each hemisphere (see *Methods*), in both OFF- and ON-levodopa conditions. For each burst, we quantified conventional time-frequency (TF) features (i.e. duration, amplitude, peak frequency, frequency span) as well as overall burst rate. The group-level analysis, on the one hand, showed small but significant increases in burst amplitude (Figures 1b, 2b), peak frequency (Figures 1c, 2c), and frequency span (Figures 1d, 2d) after levodopa administration, in both the cortex and STN. However, overall burst rates did not differ significantly between ON and OFF conditions in either cortex (Figure 1e,f; *M_OFF_*= 3.771 Hz, *SD* = 0.418 Hz, *M_ON_* =3.593 Hz, *SD* = 0.371 Hz, *p* = 0.136) or the STN (Figure 2e,f; *M_OFF_* = 3.890 Hz, *SD* = 0.447 Hz, *M_ON_* = 3.673 Hz, *SD* = 0.292 Hz, *p* = 0.193). On the other hand, TF features showed high inter-subject variability at the individual level. In the cortex, burst duration, amplitude, and peak frequency each increased, decreased, or remained unchanged with levodopa depending on the patient (Tables S2-S4). A similar pattern was observed in the STN, with a modest tendency toward increased burst amplitude in the ON-medication condition (Table S7), but no consistent effects on duration or peak frequency (Tables S6, S8). Frequency span was unaffected by medication in both regions (Table S5, S9). In summary, while levodopa induced subtle modulations of time-frequency features at the group level, these effects were inconsistent across individuals and not reflected in overall burst rate, suggesting that conventional metrics may lack the sensitivity required to track medication effects reliably.

**Figure 1.**
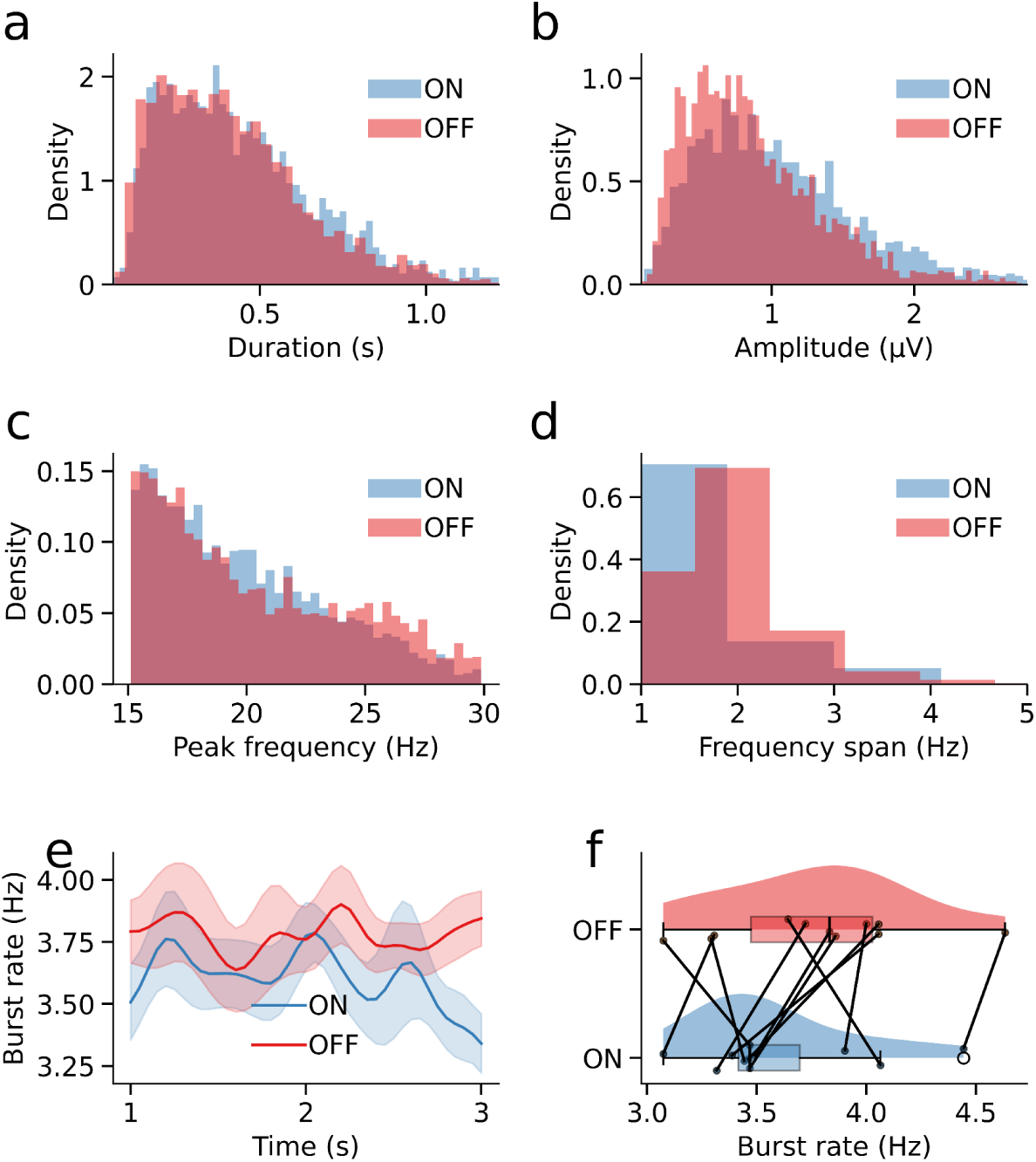
Time frequency-based features and overall rate of sensorimotor cortical bursts. Duration (a), amplitude (b), peak frequency (c), and frequency span (d) of sensorimotor cortical bursts on and off medication. e) Mean overall burst rate (shaded area indicates standard error) across patients off and on medication over time. f) Mean overall burst rate across the entire epoch. Points represent individual subjects, and black lines connect data points from the same subject.

**Figure 2.**
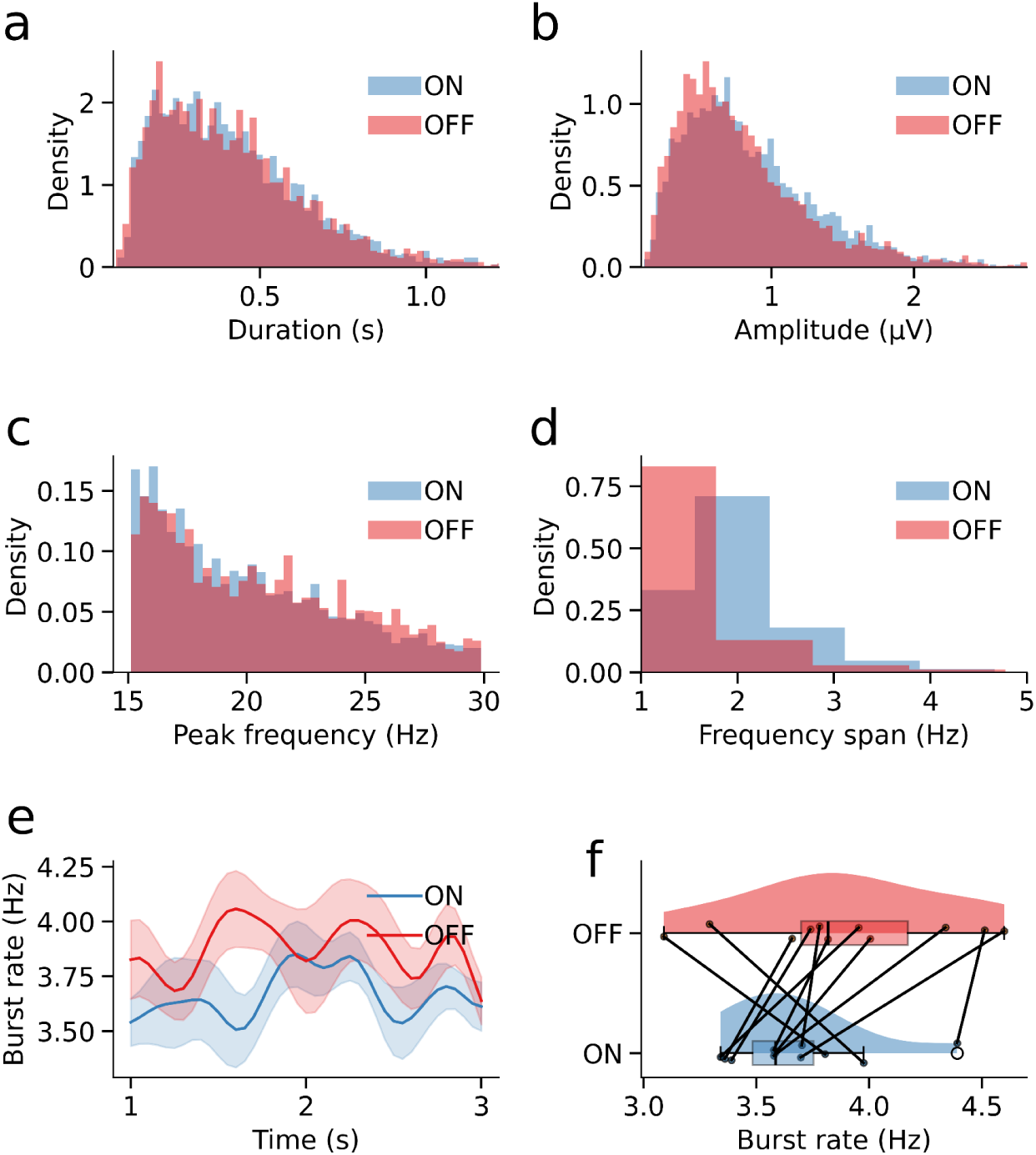
Time frequency-based features and overall rate of STN bursts. Duration (a), amplitude (b), peak frequency (c), and frequency span (d) of STN bursts on and off medication. e) Mean overall burst rate (shaded area indicates standard error) across patients off and on medication over time. f) Mean overall burst rate across the entire epoch. Points represent individual subjects, and black lines connect data points from the same subject.

### Bursts in the sensorimotor cortex and STN have similar waveforms

Because time-frequency-based beta burst features are derived from time-frequency decomposition of the burst event time series, and the correspondence between temporal and time-frequency-based burst features is not one-to-one (Jones, 2016; Szul et al., 2023), we then focused our analysis on burst waveform shapes. In line with previous work, we found that sensorimotor cortical bursts, both OFF- and ON-levodopa, had a wavelet-like median waveform shape with a strong central negative deflection and surrounding positive deflections (Figure 3a-c; Sherman et al., 2016; Bonaiuto et al., 2021; Brady and Bardouille, 2022; Rayson et al., 2023; Szul et al., 2023). Interestingly, STN bursts had a very similar median waveform shape (Figure 3d-f). Also in line with previous work, individual burst waveforms exhibited a large amount of variance around the median (Rayson et al., 2023; Szul et al., 2023). Both cortical and subcortical bursts exhibited slight increases in median waveform amplitude in the ON-levodopa state compared to the OFF-levodopa condition (Figure 3c,f), but also increases in waveform variability around the median (Figure 3b,e).

**Figure 3.**
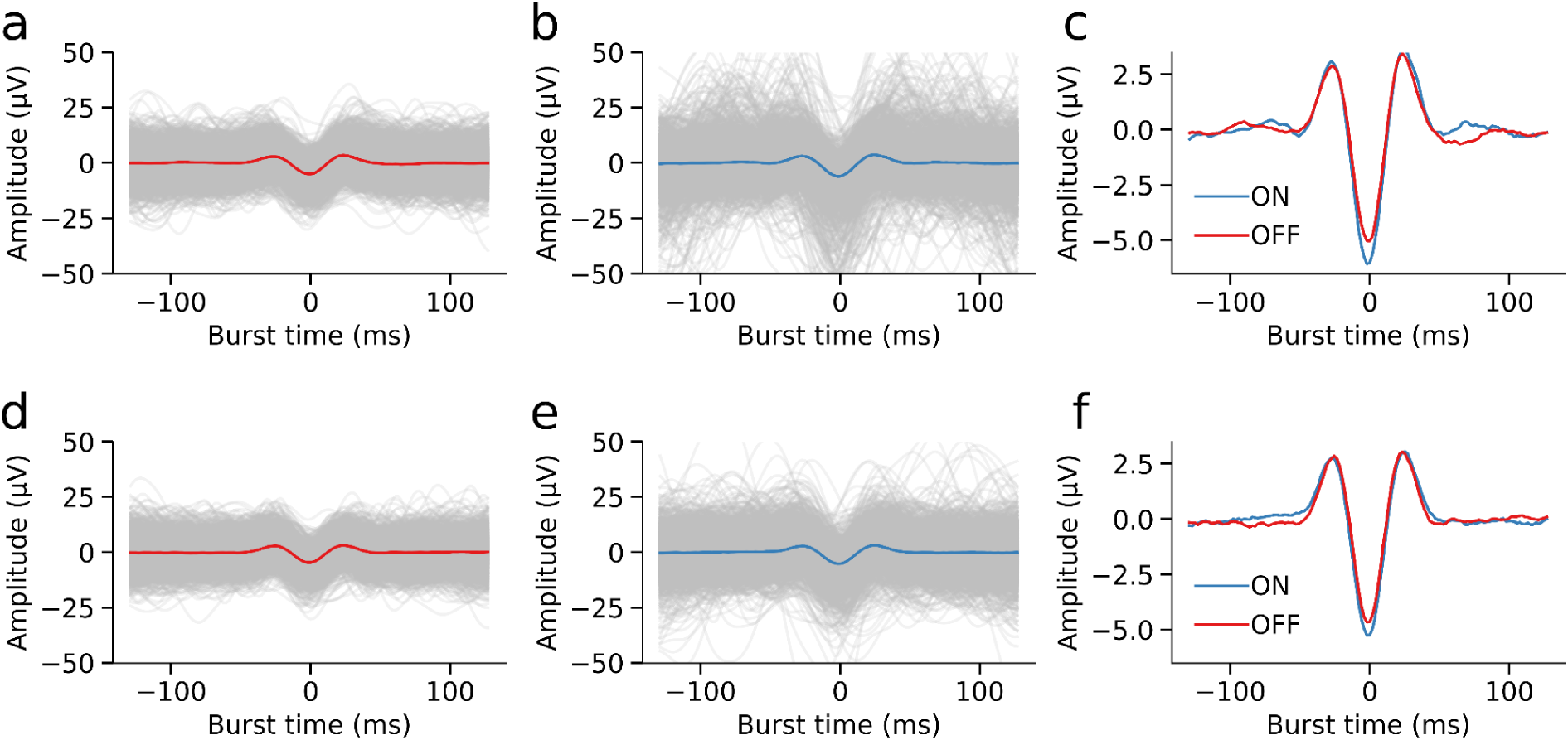
Cortical (a, b, c) and STN (d,e,f) burst waveforms. a) Waveform of each detected sensorimotor cortical burst OFF-levodopa. The red line denotes the median waveform. b) As in a, for sensorimotor cortical bursts detected ON-levodopa. The blue line represents the median waveform. c) Median sensorimotor cortical burst waveforms ON- and OFF-levodopa. d-f) As in a-c, for STN bursts.

It has been suggested that variability in burst waveform shape may reflect variability in function, and principal component analysis (PCA) has been used to identify burst waveform motifs that are differentially rate-modulated before, during, and after movement (Rayson et al., 2023; Szul et al., 2023). Therefore, we then examined how waveform variability changes between the OFF- and ON-levodopa conditions. To determine if sensorimotor cortical bursts and STN bursts had the same pattern of waveform variability, we ran separate PCAs on burst waveforms from each region and compared the resulting eigenvectors. Most principal components (PCs) were highly similar across regions (Spearman’s *ρ* = 0.52-0.99; Figure 4). For nearly all PCs, the most similar component from the sensorimotor cortical PCA was the corresponding one of the STN PCA, indicating that the waveform motif and relative percentage of variance explained was very similar across these regions.

**Figure 4.**
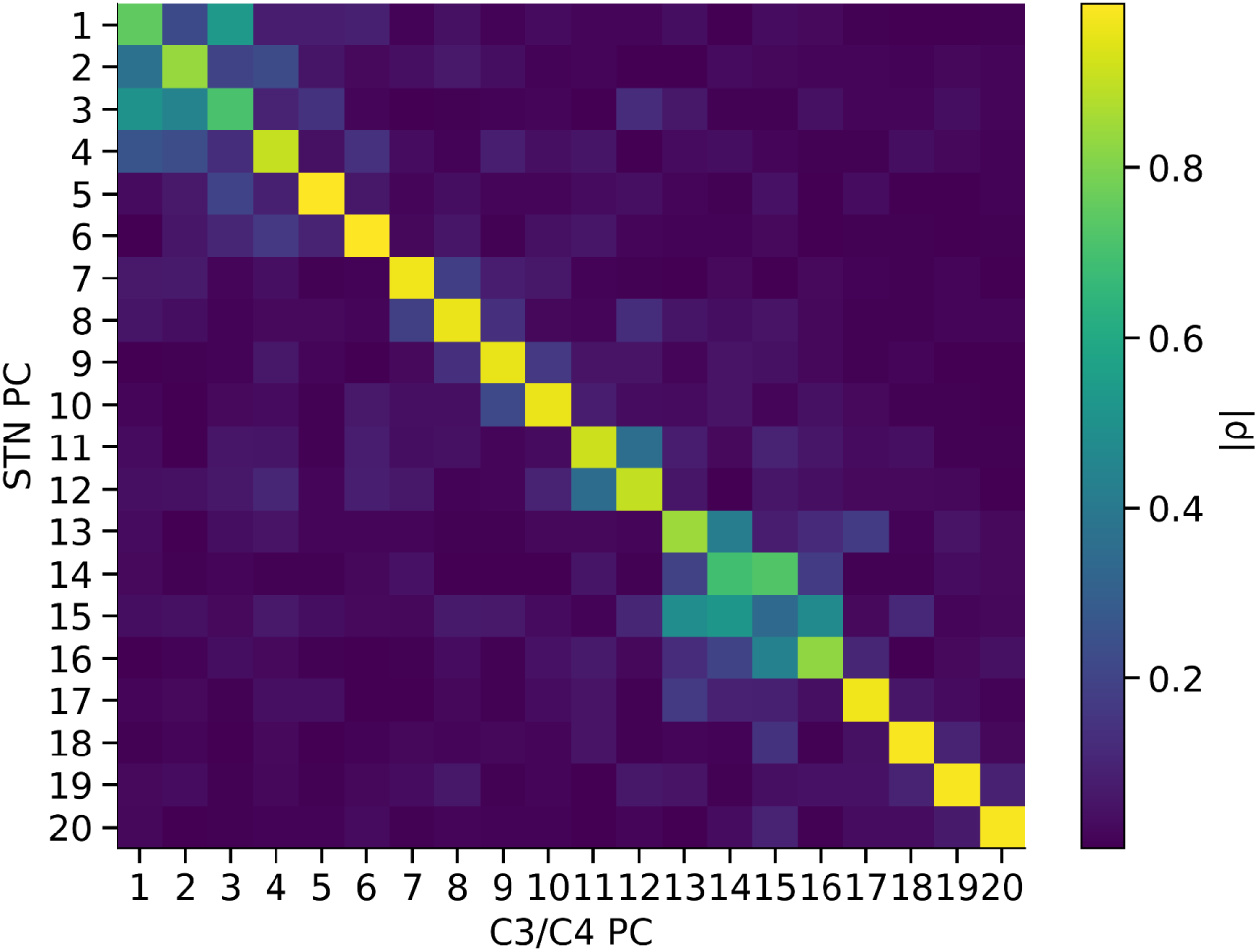
Sensorimotor cortical bursts and STN bursts have similar patterns of waveform variability. Absolute value of the correlation coefficient between eigenvectors from the PCA ran only on sensorimotor cortical bursts, and those from the PCA ran only on STN bursts.

### PCA reveals a variety of burst waveform motifs modulated by levodopa

Given the similarity in both waveform shape and variability between cortical and subcortical bursts, we then applied a single global PCA to all burst waveforms, allowing us to characterize them within a shared low-dimensional space. Components 1-12 were retained for further analysis based on a permutation test using temporally shuffled waveforms, identifying those that captured significantly more variance than expected by chance (*p* < 0.001; Figure 5a). To identify waveform motifs modulated by levodopa, we computed the difference in mean PC loadings between OFF- and ON-medication conditions for each patient and component. A second permutation test, in which condition labels were shuffled, revealed significant levodopa-induced shifts in components 1-2, 6-7, and 9-11 (*p* < 0.001). These components captured specific deviations from the median waveform, such as alterations in the central negative deflection, surrounding peaks, or more peripheral features (Figure 5b). These results were robust to the choice of STN contacts, with similar findings when including all contacts (Figure S1). Because changes in mean PC loading are not a direct reflection of shifts in the prevalence of bursts with different waveform types, we next examined levodopa-induced changes in waveform-specific burst rate, stratified by PC loading.

**Figure 5.**
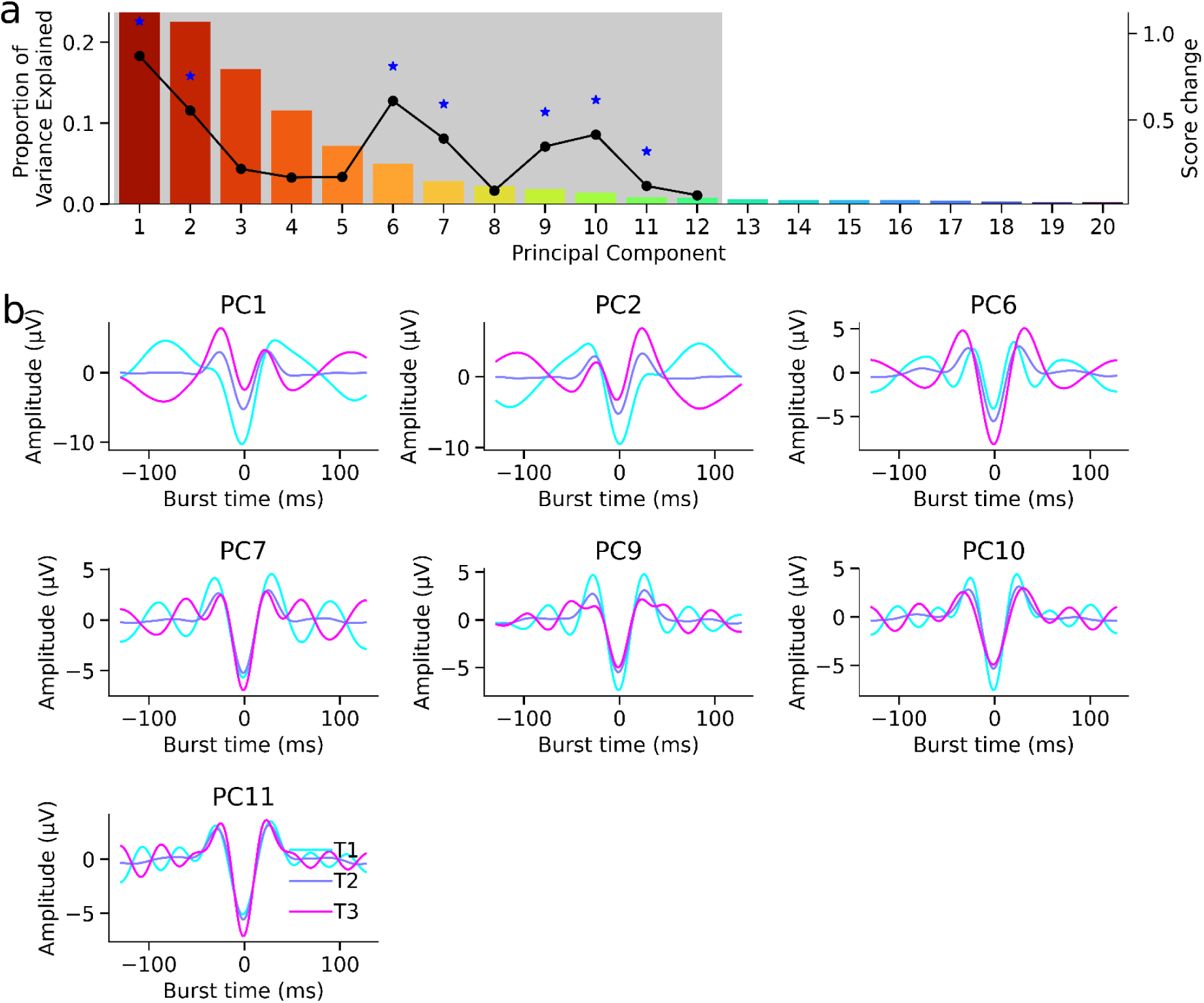
PCA reveals distinct beta burst waveform motifs modulated by levodopa. a) Histogram showing the proportion of the variance explained by each principal component (PC). Grey shading denotes components that explained significantly more variance than expected by chance, based on a temporal shuffling permutation test. The black line indicates the mean absolute change in PC loading between OFF- and ON-levodopa conditions, averaged across patients. Blue stars mark the PCs with significant medication-related changes in mean loading. b) For each significant PC, mean burst waveform shapes of bursts in the first (cyan), second (blue), and third (magenta) tertile of the loading distribution along that component, illustrating the waveform motifs captured by each dimension.

### Levodopa modulates the rate of sensorimotor cortex bursts with specific waveform shapes

For each PC that showed levodopa-induced modulation of burst waveform shape, we stratified bursts into tertiles based on their loading along that component, and computed burst rates within each tertile across time. Permutation tests were used to assess significant differences in waveform-specific burst rates between OFF- and ON-levodopa conditions. In the sensorimotor cortex, levodopa induced consistent, waveform motif-specific rate changes across patients. Notably, for PC9, bursts in the first tertile (reflecting one end of the waveform continuum) significantly increased in rate with medication (*M_OFF_* = 1.15 Hz, *SD* = 0.48 Hz; *M_ON_* = 1.51 Hz, *SD* = 0.40 Hz; *p* = 0.033; Figure 6e), while bursts in the third tertile significantly decreased (*M_OFF_* = 1.38 Hz, *SD* = 0.50 Hz; *M_ON_* = 0.96 Hz, *SD* = 0.30 Hz; *p* = 0.03; Figure 6g). The rate of intermediate bursts (second tertile) was unchanged (*M_OFF_* = 1.23 Hz, *SD* = 0.31 Hz; *M_ON_* = 1.10 Hz, *SD* = 0.39 Hz; *p* = 0.582; Figure 6f). No PC showed a consistent rate modulation in the STN. These findings were robust to the choice of subthalamic contact, with similar results obtained when using all STN contacts (Figure S2). Together, these results indicate that levodopa induces a specific redistribution of sensorimotor cortex beta bursts along the waveform axis defined by PC9 (Figure 6c,d), rather than a uniform change in burst rate. However, none of these levodopa-induced changes in waveform-specific sensorimotor cortex burst rates were correlated with clinical improvement.

**Figure 6.**
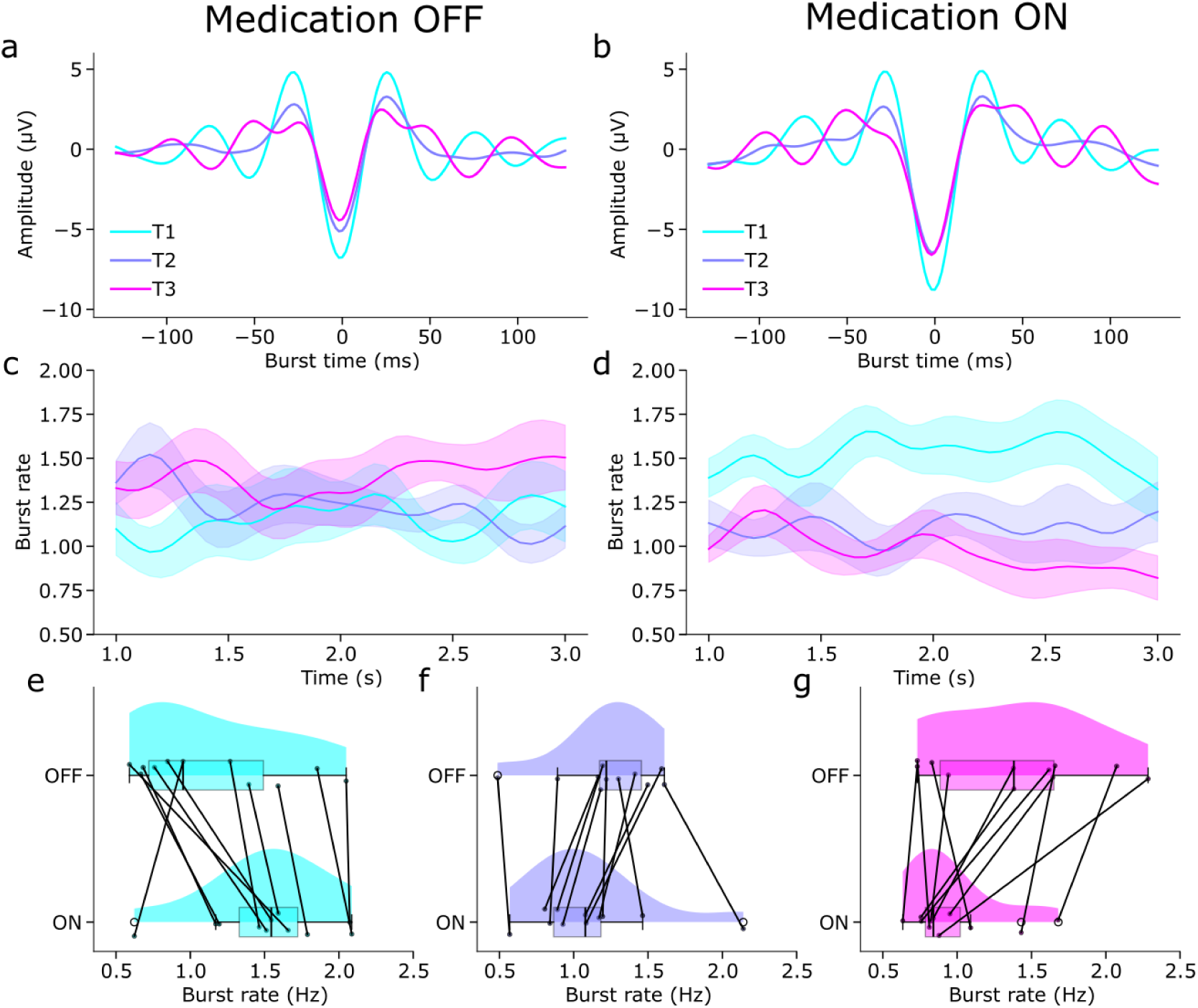
Medication induces a consistent shift in waveform-specific sensorimotor cortex burst rates along PC9. a) Mean burst waveform shapes of sensorimotor cortex bursts during medication OFF with a score within the first (cyan), second (blue), and third (magenta) tertile of all burst scores for PC9. b) As in a, for bursts during medication ON. c) Mean smoothed burst rate (shaded area indicates standard error) across patients off medication over time for each tertile of PC9. d) As in c, during medication ON. e-g) Mean burst rate across the entire epoch for the first (e), second (f), and third (g) tertiles of PC9. Points represent individual subjects and black lines connect data points from the same subject.

### Levodopa-induced increases in burst waveform-specific functional connectivity between the STN and sensorimotor cortex predict clinical improvement

Given the effects of levodopa on beta burst dynamics in the sensorimotor cortex, we next asked whether levodopa modulates the functional connectivity (FC) between beta bursts in the STN and sensorimotor cortex. We computed signed Granger causality between bursts stratified by tertiles of PC9 loadings across C3, C4, left STN, and right STN using a negative binomial generalized linear model (GLM) framework (Kim et al., 2011). In this approach, Granger causality quantifies how the occurrence bursts of specific waveform shapes at one site predict the probability of specific kinds of bursts appearing shortly afterward at another site. Because our burst detection algorithm cannot detect overlapping bursts, we focused the FC analyses on connections between regions. Whereas FC within and between left STN and right STN was minimal in both medication states, with a small amount of connectivity from left to right STN in the OFF-medication and not in the ON-medication condition (Figure 7a,b), levodopa increased FC between the STN and sensorimotor cortex in a waveform-specific manner. Notably, levodopa-induced increases in FC between the sensorimotor cortex and STN were largely intra-hemispheric, whereas for the STN itself, they occurred between hemispheres (i.e., between the right-STN and left-STN). In the ON-medication condition, we observed stronger connectivity from both C3 and C4 to the left and right STN contacts, and vice versa, particularly involving bursts in the first and third tertiles of PC9. Connectivity between C3 and C4 was reduced in the ON-medication condition and limited to these same tertiles. Permutation tests confirmed significant levodopa-induced increases in FC from third-tertile bursts in C3 to third-tertile bursts in left STN, and from first-tertile bursts in C4 to first-tertile bursts in right STN, consistent with prior evidence that cortical bursts precede and potentially drive STN bursts (Figure 7c; Yao et al., 2025). Levodopa therefore selectively enhances functional connectivity between STN and sensorimotor cortex during beta bursts of specific waveform motifs defined by PC9.

**Figure 7.**
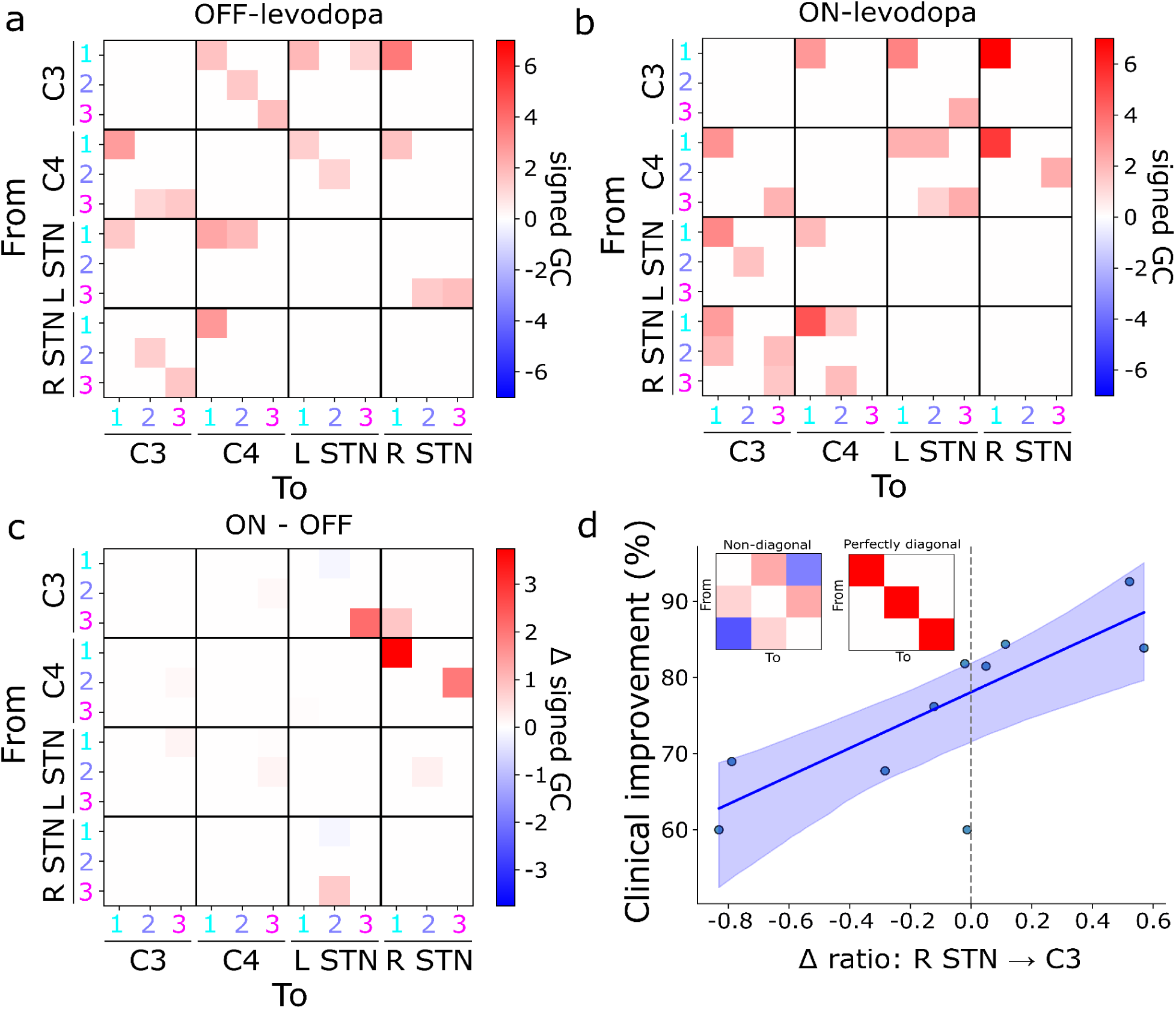
Levodopa increases reciprocal functional connectivity between sensorimotor cortex and STN, specifically during bursts of specific waveform shapes. a) Significant Granger causality between bursts in each tertile of PC9 in left and right hemisphere sensorimotor cortex and STN OFF-levodopa. Horizontal and vertical lines separate different regions. b) As in a, ON-levodopa. c) Significant differences in Granger causality between conditions (ON- minus OFF-levodopa). d) An increase in the diagonality in the pattern of connectivity between right STN and left sensorimotor cortex predicts clinical improvement. The solid line is the fitted linear model. The shaded region denotes the 95% confidence interval. Each dot is the data point of a single subject. The inset shows examples of sub-matrices (tertile × tertile) with lower versus higher diagonality.

We then asked whether levodopa-induced changes in beta burst waveform-specific FC between the STN and sensorimotor cortex were related to clinical improvement, defined as the percentage change in UPDRS III score (Tinkhauser et al., 2017b). To assess this, we examined FC between each tertile of one region and each tertile of another. Interestingly, changes in FC between the sensorimotor cortex and the STN for first-tertile bursts were negatively correlated with clinical improvement; whereas, for third-tertile bursts, there was a positive correlation (Figure S3.a-b). This dissociation mirrors the opposing effects of levodopa on burst rates in sensorimotor cortex: the rate of first-tertile bursts increased, whereas that of third-tertile bursts decreased. Together, these results suggest that third-tertile bursts, although less frequent under medication, are more functionally relevant within the sensorimotor-STN circuit in the ON-levodopa condition.

In order to determine if FC during bursts of the same shape relates to clinical improvement, we computed the trace-to-spectral norm ratio of the 3×3 Granger causality submatrix (tertile × tertile) linking each pair of regions (C3, C4, left STN, right STN) in the ON and OFF-medication conditions. This metric quantifies how close the connectivity matrix is to being diagonal, reflecting the extent to which bursts of a given waveform class influence bursts of similar shape in a target region. We then tested whether the medication-induced change in this metric for each region pair predicted clinical improvement across subjects. A significant positive relationship was observed for the connection from right STN to C3 (Spearman’s *ρ* = 0.78, *p* = 0.011; robust slope = 18.38, *p* = 0.010; Figure 7d). Clinical improvement was therefore specifically related to levodopa-induced changes in the functional connectivity from STN to sensorimotor cortex within a particular waveform motif, introducing a new waveform-based metric for evaluating medication effects.

## Discussion

Our findings suggest that levodopa modulates beta burst waveform motifs in both the STN and the sensorimotor cortex. Remarkably, specific waveform motifs in the sensorimotor cortex were selectively rate-modulated by levodopa. Furthermore, levodopa increased burst waveform-specific functional connectivity between the STN and the sensorimotor cortex, and this modulation was correlated with levodopa-induced clinical improvement. Therefore, our study establishes a precise framework for characterizing PD-related beta dynamics at the network level, providing new patient-specific biomarker candidates for both disease severity and treatment response.

We first examined the local dynamics of beta bursts in both the STN and sensorimotor cortex both before and after levodopa administration, focusing on TF features such as power amplitude, peak frequency, duration, frequency span, and overall burst rates. However, at the individual subject level, these features showed inconsistent medication effects. We then analyzed beta burst waveforms in the time domain, revealing that both cortical and subcortical bursts share a characteristic wavelet-like average shape, consistent with prior findings in the sensorimotor cortex (Sherman et al., 2016; Bonaiuto et al., 2021; Brady and Bardouille, 2022; Rayson et al., 2023; Szul et al., 2023). Despite this similarity, individual burst waveforms exhibited a large amount of variance around the median, reinforcing previous observations that beta bursts deviate significantly from their average waveform (Howe et al., 2011; Karvat et al., 2020; Kosciessa et al., 2020; Yeh et al., 2020; Bonaiuto et al., 2021; Szul et al., 2023). Notably, this waveform diversity is not captured by standard TF-based analyses (Szul et al., 2023) suggesting that bursts with similar TF signatures may reflect distinct neural processes. Such heterogeneity may underlie the diverse functional roles attributed to beta activity (Pfurtscheller et al., 1997; Salenius and Hari, 2003; Engel and Fries, 2010; Kilavik et al., 2013; Little and Brown, 2014; Reuter et al., 2022). A promising avenue for future work is to investigate this variability in STN beta burst waveforms through computational modeling, as has been done for beta burst generation in the somatosensory cortex (Jones et al., 2009; Sherman et al., 2016; Law et al., 2022).

We applied PCA to classify the diversity of beta burst waveforms in both cortical and subcortical regions, identifying the principal dimensions that capture waveform variability. This analysis revealed a range of distinct beta burst motifs, with similar patterns of variability across the sensorimotor cortex and STN. Notably, levodopa selectively modulated specific waveform motifs in both regions, but only in the sensorimotor cortex were these medication-induced changes in burst rates consistent across patients. Furthermore, these levodopa-induced changes were accompanied by selective increases in functional connectivity between the STN and sensorimotor cortex during bursts with specific waveform motifs, as quantified by Granger causality analyses. This connectivity was most prominent between bursts of similar waveform classes across regions, suggesting that levodopa enhances motif-aligned communication rather than uniformly increasing coupling. This modulation was linked to levodopa-induced clinical improvement, specifically through increased diagonality of the connectivity matrix between right STN and left sensorimotor cortex. This suggests that clinical improvement is associated with more selective, waveform-specific interactions within cortico-basal ganglia circuits. These results introduce burst waveform-specific functional connectivity between the STN and sensorimotor cortex as a precise, waveform-based metric for assessing medication effects and underscore the need for targeted analysis of distinct burst motifs in future studies of PD.

Interestingly, our analyses revealed that levodopa can simultaneously increase the rate of certain sensorimotor cortex burst shapes (bursts in the first tertile of PC9) while reducing others (those in the third tertile). Despite this decrease in third tertile bursts, we found that increases in FC between the sensorimotor cortex and STN involving third tertile bursts predicted greater clinical improvement during medication. This suggests that levodopa enhances the burst-mediated effective information transfer, in the Granger causality sense, across the sensorimotor-STN circuitry, despite a reduction in burst occurrence. This is consistent with previous research on aperiodic bursts (i.e., neuronal avalanches), which demonstrated that while levodopa reduces the number of avalanches, they tend to spread more in the ON-levodopa condition (Agouram et al., 2025). Hence, these findings suggest that the dynamics of beta burst waveforms not only mirror local neural processes but also play a role in broader cortico-basal ganglia interactions. Moreover, this apparent normalization of burst dynamics aligns with the idea that balancing distinct burst types is beneficial for motor performance (Tinkhauser et al., 2017b). In other words, medication may refine the network’s reliance on more adaptive bursts while pruning pathological burst activity, creating fewer third tertile events overall but rendering them more functionally relevant.

Previous studies have linked beta burst features to motor impairment (Anidi et al., 2018; Lofredi et al., 2019; Kehnemouyi et al., 2021), medication responses (Tinkhauser et al., 2017b; Duchet et al., 2021), and deep brain stimulation effects (Tinkhauser et al., 2017a; Schmidt et al., 2020; Pauls et al., 2022). However, these metrics have predominantly relied on TF decompositions or band-pass filtered signal envelopes. In contrast, prior research has shown that TF-based burst features do not effectively capture differences in waveform structure in the temporal domain (Jones, 2016; Szul et al., 2023), a limitation that aligns with our findings. Waveform-based measures may offer greater sensitivity for early PD diagnosis compared to more coarse TF-based burst features. Specific burst motifs could be rapidly identified and targeted by DBS systems to optimize stimulation timing. Time domain template matching (Papadopoulos et al., 2024) could further reduce detection-to-stimulation delays, potentially improving treatment efficacy. By shifting the focus from aggregate beta activity to distinct waveform motifs, this approach has the potential to refine both diagnostic tools and neuromodulatory therapies for PD (Szul et al., 2023).

More generally, these results prompt a reappraisal of the notion that beta bursts are generated in the cortex by converging synaptic drives that do not themselves resemble beta bursts (Sherman et al., 2016; Szul et al., 2023). The similarity between cortical and subcortical bursts, not only in terms of median waveform shape, but also variability around the median, and the reciprocal functional connectivity between sensorimotor cortex and STN suggests that cortical beta bursts are generated via recurrent interactions between cortex and basal ganglia. Moreover, the fact that this connectivity was specific to bursts with particular waveforms suggests the exciting possibility that cortical beta bursts recorded via M/EEG provide a window for measuring neuronal activity in the basal ganglia at a temporal resolution unobtainable with other non-invasive methods.

## Methods

### Participants, Surgical Procedures, Experiments, and Data Collection

This study utilized resting-state data from PD patients, partially overlapping with the dataset used by Tinkhauser et al. (2018), which consists of 11 patients diagnosed with advanced Parkinson’s disease who received deep brain stimulation (DBS) therapy targeting the subthalamic nucleus (STN). Neural recordings were obtained using DBS leads and EEG electrodes positioned bilaterally over motor regions. Data collection occurred under two conditions: OFF-levodopa medication (prior to administration) and ON-levodopa medication (post-administration). The medication conditions were systematically balanced for the cohort. Recordings in the ON-medication condition were done 45 minutes following levodopa administration. The ON-state was confirmed by clinical evaluation using the MDS-Unified Parkinson’s Disease Rating Scale (MDS-UPDRS).

Ten patients’ detailed clinical data are included in the supplementary part (Table S1); clinical data for one patient were not available. This work was approved ethically by the local ethics committee (Comité de Protection des Personnes (CPP) Sud Méditerranée I) (registration number RCB: 2009-A00913-54), and all participants gave written informed consent. Medtronic 3389 DBS leads were used during bilateral DBS surgery. To guarantee accurate lead placement during surgery, intraoperative micro-recordings and macro-stimulation were used. Following surgery, validation was achieved by combining fused preoperative MRI and postoperative CT scans. As part of the experimental protocol, the DBS electrodes were temporarily externalized before being connected to the implantable pulse generator about five days later. For the OFF-dopa state, patients did not take any dopaminergic medications the night before recordings. For the ON-state, patients received a dose of oral levodopa in a dispersible form equivalent to 150% of their usual morning dose (levodopa challenge). The recordings started 30 minutes following the intake of the levodopa dose and the switch to the ON-state was confirmed clinically using the UPDRSIII. Neural signals were recorded while patients were seated comfortably at rest for a duration of two to three minutes. Local field potentials (LFPs) were recorded simultaneously from eight contacts in the STN (four in each hemisphere), along with scalp EEG signals from six channels placed over the bilateral sensorimotor areas (F3, Fz, F4, C3, C4, Cz). We used the Spike2 Software to minimize and eliminate spike artifacts. The data, which ranged from 119.2 to 155.5 seconds in duration (mean: 127.2 ± 2.5 seconds), was imported into Matlab.

### Data Preprocessing

The Fieldtrip toolbox was used for signal preprocessing (Oostenveld et al., 2011). The continuous EEG signal was first filtered using a high-pass filter at 1.3 Hz and a low-pass filter at 45 Hz using a two-pass Butterworth filter and a Hamming window. The data was then resampled to 512 Hz and split into 4-second segments. After that, the data was examined visually to find and eliminate any noisy segments.

### Beta Burst Detection Algorithm

Beta bursts were identified through a new adaptive burst detection method (Rayson et al., 2023; Szul et al., 2023), which was created to identify all potential burst events across a broad range of beta amplitudes, without the use of a fixed, global threshold. The algorithm iteratively fits a 2D Gaussian to the global maxima for each trial’s TF decompositions, subtracting the fitted Gaussian from the TF decomposition, and proceeding with the resulting global maxima, continuing until no additional bursts are found in the trial. The superlet transform (Moca et al., 2021) was used to perform TF decomposition. This transform offers an optimal balance between time and frequency resolution, making it appropriate for identifying brief bursts within specific frequency bands. An adaptive superlet transform using Morlet wavelets was applied, with varying central frequencies (1.5 - 40 Hz) and a fixed number of cycles (4 cycles) within a Gaussian envelope. The wavelet order, which is the multiplier for the number of cycles in the wavelet, varied linearly from 1 to 30 over the frequency range. The TF decomposition was used to compute the power spectral density (PSD) for each channel by averaging the TF power over time for individual trials and subsequently across trials. The PSD for each channel was then parameterized using the specparam algorithm (Donoghue et al., 2020), which estimates and subtracts the aperiodic spectrum from each individual trial of TF decomposition. The burst detection process is then applied to the residual amplitude (Brady and Bardouille, 2022), which finds the global maximum in the TF space and fits a 2D Gaussian to this peak. The Gaussian parametrization characterizes burst features in the TF domain, such as peak time, duration (full width at half maximum (FWHM) in time), peak amplitude, peak frequency, and frequency span (FWHM in frequency). The Gaussian is then removed from the TF decomposition, and the next iteration proceeds with the resulting residual TF matrix. The process stops when no peaks exceed the noise floor, which was recalculated in each iteration and was determined as two standard deviations above the average power across all time and frequency bins. To minimize edge effects, detection was limited to TF data in the 10-33 Hz range, with bursts of peak frequencies within the beta band (13-30 Hz) maintained for further analysis. For more information, refer to (Szul et al., 2023).

Subsequently, the peak time was used to extract the waveform for each detected burst from the preprocessed signal, which was broadband-pass filtered between 1.3 and 45 Hz. The time window for waveform extraction was 260 ms, which was determined by the decrease in beta lagged coherence over increasing lag cycles (Szul et al., 2023). The time series within this window, centered on the peak time, was extracted. To determine the signal deflection at the peak amplitude, we aligned the burst waveforms by band-pass filtering within their detected frequency span, computed the instantaneous phase using the Hilbert transform, and re-centered the “raw” waveform (before band pass filtering) around the phase minimum that was closest to the peak time in TF space (Boto et al., 2022). If the time point deviated by more than 30 ms from the TF-detected peak time, the burst was excluded. Subsequently, the DC offset was removed from the resulting waveform.

### Burst analysis

To enhance the robustness of our findings, we first located the best STN contact with the maximum beta power (13-30 Hz) under the ON-medication condition, for each patient and hemisphere, by computing the PSDs using Welch’s method (Hann window, 256-sample segments, 7/8 overlap). Then, we used only those best STN contacts and the sensorimotor cortex channels (C3 and C4) for all of the analyses.

Next, we analyzed the distribution of beta burst features (duration, peak amplitude, peak frequency, and frequency span) at the group and individual subject levels, comparing the ON and OFF medication conditions in the C3/C4 channels and STN contacts. We assessed the statistical significance at each level using permutation tests. For the group-level, we compared the observed difference in mean burst features between the two medication conditions to a null distribution generated by 100,000 random permutations of ON and OFF labels. Whereas, at the subject-level, we used the median instead of the mean. Additionally, we determined the median and interquartile range (IQR) for each burst feature in both medication states and for each patient. Also, we compared the distribution of beta burst features across all patients between the ON and OFF medication conditions for the C3/C4 channels and STN contacts.

We compared the overall burst rates between the ON and OFF medication conditions for C3/C4 channels and STN contacts across patients. Burst rates were calculated using 50 ms time bins, normalized by trial count and duration, and smoothed with a Gaussian filter (σ = 2.5). We trimmed the first and last second from each epoch to correct for edge effects. A paired permutation test was used to evaluate statistical significance. First, we computed the observed difference in mean burst rates between the ON and OFF conditions. After that, we created a null distribution by randomly flipping the signs of the paired differences over 100,000 iterations and computing the mean of the permuted differences. The p-value was defined as the proportion of permuted differences greater than or equal to the observed difference.

To investigate changes in waveform variability and classify different burst waveform shapes between the medication-ON and medication-OFF states, we used Principal Component Analysis (PCA). This analysis, which used 20 components and was implemented using the scikit-learn toolkit (Pedregosa et al., 2011), was performed on aligned waveforms, treating each time point as a feature. The PCs create a 20-dimensional coordinate system, with the origin representing the average burst waveform. The waveform of each burst is characterized by its position in this space, determined by its score on each PC, which indicates the distance from the mean waveform along each dimension. To compare the waveform variability between bursts from the sensorimotor cortex and the STN, we first ran independent PCAs on the burst waveforms from each region and used Spearman correlation to compare their eigenvectors. The findings showed that the waveform shapes and variability patterns of cortical and subcortical bursts were very similar. Based on these similarities, we performed a unified PCA on all burst waveforms, allowing us to analyze them using a common dimensional framework.

In order to determine which PCs were able to capture significant variance in the beta burst waveform data, we used a permutation-based shuffling approach (Vieira, 2012). The variance explained by the components from the real data was compared with the variance distributions from 100 iterations of shuffled data, in which rows of each feature column (time points) were randomly permuted to eliminate temporal correlations. P-values were calculated as the proportion of shuffled PCA results with lower explained variance than the original data. After applying a Bonferroni correction (*α* = 0.0025) for multiple comparisons, PCs 1-12 were found to be significant and suitable for further analysis.

To identify the waveform motifs modulated by levodopa, we first computed the magnitude of the difference in mean burst scores between the OFF and ON conditions, which was averaged across all patients. After that, a permutation test was performed, in which a null distribution of score differences for each significant PC was created by shuffling 1,000 times the ON/OFF medication labels. To evaluate statistical significance, the observed score differences were compared with this null distribution. Levodopa-modulated PCs were defined as those whose p-values fell below a Bonferroni-corrected significance threshold. This test revealed that PCs 1-2, 6-7, and 9-11 captured significant changes in waveform shape between the two conditions (*p* < 0.001). These PCs corresponded to dimensions where the waveform shape deviated from the median waveform, such as changes in the central negative deflection, surrounding peaks, or additional peripheral peaks. Since changes in the mean PC score might be caused by variations in burst rate rather than waveform shape alone, we subsequently studied the PCs in relation to waveform-specific burst rates to clarify their significance.

For each PC dimension reflecting levodopa-induced changes in waveform shape, bursts were sorted into tertiles according to their scores along that dimension and their timing within the trial. Burst rates for each tertile and condition (OFF and ON) were calculated by binning burst peak times into 50 ms time bins, normalizing by the number of trials, and smoothing with a Gaussian filter. To evaluate statistical significance, a paired permutation test was used, followed by Bonferroni correction for multiple comparisons.

### Granger causality Analysis

After identifying levodopa-induced changes in beta bursts aligned with the waveform motif defined by PC9, we next assessed whether dopaminergic state modulated functional connectivity between the sensorimotor cortex and STN during these bursts. Bursts were binned by time (bin size of 5 ms) within each tertile and region of interest (C3, C4, left STN, right STN), producing event matrices of burst counts across trials and time bins. For each subject and condition, we estimated functional connectivity using Granger causality (GC) computed from a negative binomial generalized linear model (GLM), which was adapted from (Kim et al., 2011). To determine the optimal time lag for each source-target pair, we performed 10-fold cross-validation across lags from 1 to 10 bins (5 to 50 ms), selecting the lag that maximized held-out log-likelihood. GC scores between source-target pairs were computed by comparing the log-likelihoods of full and reduced models, with reduced models obtained by removing the source from the data matrix. Signed GC values were derived by multiplying the GC score by the sign of the source coefficient in the full model. All models were conditioned on activity from the remaining units to control for shared inputs.

Because our burst detection algorithm cannot detect overlapping bursts, we did not analyze within-region connections and focused exclusively on inter-regional connectivity. To assess whether functional connectivity was significantly greater than expected by chance, we first computed a null distribution of GC values by randomly shuffling trials independently for each tertile and site, thereby preserving burst statistics while disrupting temporal relationships between regions. For each subject, this yielded a null GC matrix for each of 1,000 permutations; we then applied false discovery rate (FDR) correction across all source-target pairs to identify statistically significant connections in each condition (Figure 7a,b). To assess whether connectivity changed significantly between the ON- and OFF-medication conditions, we then performed a second, repeated-measures permutation test in which ON/OFF labels were randomly flipped within subjects. This test, repeated 10,000 times for each source-target pair, provided a null distribution against which to evaluate levodopa-induced changes in GC (Figure 7c).

### Clinical correlation analysis

To examine whether levodopa-induced changes in beta burst connectivity were related to therapeutic benefit after levodopa administration, we analyzed correlations between clinical improvement and motif-specific functional connectivity in the sensorimotor-STN circuit along PC9. To evaluate this relationship, we analyzed both individual connections (each tertile-to-tertile pair across regions) and a summary metric of connectivity diagonality (trace-to-spectral norm ratio) for each region pair (left and right STN, C3, C4). For each subject and each region pair, we calculated the relative change in this ratio between ON and OFF medication states. Clinical improvement was defined as the percentage reduction in UPDRS-III motor scores from OFF to ON medication (Tinkhauser et al., 2017b):

Clinical_improvement = ((updrs_III_off - updrs_III_on) / updrs_III_off)*100

UPDRS_III_off represents the Unified Parkinson’s Disease Rating Scale Part III score prior to medication intake, while UPDRS_III_on represents the score following medication intake.

To evaluate the relationship between clinical improvement and the change in connectivity, we used robust linear regression and Spearman’s rank correlation for each connection. The regression model was estimated using M-estimators, and the significance of the slope was assessed via a non-parametric permutation test (*N* = 10,000). Confidence intervals around the regression line were obtained by bootstrapping (*N* = 10,000). FDR correction was used to account for multiple comparisons.

## Data availability and code access

Due to the sensitive clinical nature of the data, it cannot be made publicly available. The code used in this study is available at: https://github.com/danclab/beta_bursts_ldopa.

## Data Availability

Due to the sensitive clinical nature of the data, it cannot be made publicly available. The code used in this study is available at:https://github.com/danclab/beta_bursts_ldopa

## Supplementary Material

**Table S1:** Clinical data from 10 patients. F = female; M = male; L = left; R = right; AR = Akinetic rigid; UPDRS = Unified Parkinson’s Disease Rating Scale. Clinical data for s15 could not be recovered.

| Patient | Age range (years) | Sex | Disease duration (years) | First symptom | Side onset | UPDRS III OFF | UPDRS III ON | Delay between surgery and recording (days) | Medication |
| --- | --- | --- | --- | --- | --- | --- | --- | --- | --- |
| s02 | 51-55 | M | 14 | tremor | R | 27 | 5 | 3 | ropinirole 8mg; l-dopa 1250mg/day; entacapone; rasagiline |
| s04 | 66-70 | F | 12 | AR | R | 31 | 10 | 3 | ropinirole 8mg; l-dopa 700mg/day; entacapone |
| s05 | 66-70 | M | 10 | tremor | R | 32 | 5 | 3 | piribedil 150mg/day; l-dopa 1250mg/day; entacapone |
| s06 | 51-55 | F | 8 | tremor | L | 40 | 16 | 3 |  |
| s07 | 51-55 | F | 10 | AR | L | 11 | 2 | 3 | ropinirole 16mg; l-dopa 400mg |
| s08 | 61-65 | F | 12 | AR | L | 21 | 5 | 3 | ropinirole 20mg; l-dopa 1200mg; entacapone |
| s10 | 56-60 | M | 17 | AR | R | 27 | 2 | 3 | pramipexole 2.1mg; l-dopa 1200mg; entacapone |
| s16 | 61-65 | M | 14 | tremor | L | 29 | 9 | 3 | piribedil 150mg; l-dopa 900mg; entacapone |
| s17 | 61-65 | M | 7 | tremor | L | 31 | 5 | 3 | ropinirole 24mg; l-dopa 800mg; entacapone; rasagiline 1mg |
| s18 | 56-60 | F | 8 | tremor | L | 20 | 8 | 5 | piribedil 150mg; l-dopa 1000mg; entacapone |

**Table S2:** Sensorimotor cortical burst duration.

| Subject | Off medication (median: IQR) | On medication (median: IQR) | Statistic | Change |
| --- | --- | --- | --- | --- |
| s02 | 402.34 ms (274.41-550.78 ms) | 460.94 ms (304.69-591.80 ms) | p=0.004 | increase |
| s04 | 382.81 ms (237.30-554.69 ms) | 449.22 ms (285.16-636.72 ms) | p=0.006 | increase |
| s05 | 355.47 ms (242.19-500.00 ms) | 460.94 ms (296.88-634.77 ms) | p<0.001 | increase |
| s06 | 357.42 ms (234.38-515.62 ms) | 384.77 ms (256.84-535.16 ms) | p=0.103 | none |
| s07 | 414.06 ms (296.88-625.00 ms) | 449.22 ms (316.41-664.06 ms) | p=0.193 | none |
| s08 | 433.59 ms (294.92-619.14 ms) | 457.03 ms (277.34-636.72 ms) | p=0.413 | none |
| s10 | 386.72 ms (226.56-558.59 ms) | 423.83 ms (270.51-655.27 ms) | p=0.088 | none |
| s15 | 300.78 ms (203.12-449.22 ms) | 304.69 ms (195.31-441.41 ms) | p=0.866 | none |
| s16 | 367.19 ms (250.00-507.81 ms) | 396.48 ms (269.53-590.82 ms) | p=0.125 | none |
| s17 | 394.53 ms (226.56-542.97 ms) | 343.75 ms (210.94-480.47 ms) | p=0.016 | decrease |
| s18 | 375.00 ms (226.56-550.78 ms) | 378.91 ms (250.00-562.50 ms) | p=0.852 | none |

**Table S3:** Sensorimotor cortical burst peak amplitude.

| Subject | Off medication (median: IQR) | On medication (median: IQR) | Statistic | Change |
| --- | --- | --- | --- | --- |
| s02 | 0.81 μV (0.60-1.21 μV) | 1.16 μV (0.81-1.62 μV) | p<0.001 | increase |
| s04 | 0.61 μV (0.41-0.82 μV) | 1.04 μV (0.75-1.39 μV) | p<0.001 | increase |
| s05 | 0.60 μV (0.43-0.81 μV) | 1.00 μV (0.71-1.30 μV) | p<0.001 | increase |
| s06 | 0.79 μV (0.55-1.10 μV) | 0.84 μV (0.55-1.17 μV) | p=0.234 | none |
| s07 | 0.94 μV (0.67-1.27 μV) | 1.30 μV (0.94-1.73 μV) | p<0.001 | increase |
| s08 | 1.51 μV (1.14-2.01 μV) | 1.44 μV (1.03-1.94 μV) | p=0.165 | none |
| s10 | 0.83 μV (0.56-1.16 μV) | 1.06 μV (0.78-1.39 μV) | p<0.001 | increase |
| s15 | 0.45 μV (0.30-0.68 μV) | 0.48 μV (0.32-0.67 μV) | p=0.109 | none |
| s16 | 0.94 μV (0.71-1.28 μV) | 0.99 μV (0.75-1.34 μV) | p=0.175 | none |
| s17 | 0.66 μV (0.46-0.91 μV) | 0.63 μV (0.44-0.87 μV) | p=0.320 | none |
| s18 | 0.85 μV (0.57-1.21 μV) | 1.19 μV (0.72-1.91 μV) | p<0.001 | increase |

**Table S4:** Sensorimotor cortical burst peak frequency.

| Subject | Off medication (median: IQR) | On medication (median: IQR) | Statistic | Change |
| --- | --- | --- | --- | --- |
| s02 | 17.44 Hz (16.28-19.78 Hz) | 17.44 Hz (15.89-20.36 Hz) | p=1.000 | none |
| s04 | 24.06 Hz (19.00-26.49 Hz) | 22.50 Hz (19.39-24.83 Hz) | p=0.002 | decrease |
| s05 | 18.61 Hz (16.28-22.11 Hz) | 18.22 Hz (16.28-20.17 Hz) | p=0.554 | none |
| s06 | 19.00 Hz (17.44-21.33 Hz) | 19.39 Hz (17.44-20.56 Hz) | p=0.450 | none |
| s07 | 18.22 Hz (16.67-20.94 Hz) | 20.56 Hz (17.06-23.67 Hz) | p<0.001 | increase |
| s08 | 18.61 Hz (16.67-21.72 Hz) | 22.11 Hz (19.39-24.83 Hz) | p<0.001 | increase |
| s10 | 16.67 Hz (15.50-18.61 Hz) | 17.06 Hz (15.60-18.22 Hz) | p=0.107 | none |
| s15 | 20.56 Hz (17.83-23.67 Hz) | 20.17 Hz (17.44-23.47 Hz) | p=0.559 | none |
| s16 | 22.50 Hz (19.39-26.00 Hz) | 17.83 Hz (16.28-20.17 Hz) | p<0.001 | decrease |
| s17 | 22.50 Hz (17.83-26.00 Hz) | 20.17 Hz (17.44-23.28 Hz) | p<0.001 | decrease |
| s18 | 17.83 Hz (16.28-19.39 Hz) | 17.44 Hz (15.89-20.94 Hz) | p=0.492 | none |

**Table S5:** Sensorimotor cortical burst frequency span.

| Subject | Off medication (median: IQR) | On medication (median: IQR) | Statistic | Change |
| --- | --- | --- | --- | --- |
| s02 | 1.56 Hz (1.56-1.56 Hz) | 1.56 Hz (1.56-1.94 Hz) | p=1.000 | none |
| s04 | 1.56 Hz (0.78-1.56 Hz) | 1.56 Hz (1.56-2.33 Hz) | p=1.000 | none |
| s05 | 1.56 Hz (0.78-1.56 Hz) | 1.56 Hz (1.56-2.33 Hz) | p=1.000 | none |
| s06 | 1.56 Hz (0.78-1.56 Hz) | 1.56 Hz (0.78-1.56 Hz) | p=1.000 | none |
| s07 | 1.56 Hz (1.56-1.56 Hz) | 1.56 Hz (1.56-2.33 Hz) | p=1.000 | none |
| s08 | 1.56 Hz (1.56-1.75 Hz) | 1.56 Hz (1.56-1.56 Hz) | p=1.000 | none |
| s10 | 1.56 Hz (0.78-1.56 Hz) | 1.56 Hz (1.56-1.56 Hz) | p=1.000 | none |
| s15 | 1.56 Hz (0.78-1.56 Hz) | 1.56 Hz (0.78-1.56 Hz) | p=1.000 | none |
| s16 | 1.56 Hz (0.78-1.56 Hz) | 1.56 Hz (1.56-1.56 Hz) | p=1.000 | none |
| s17 | 1.56 Hz (0.78-1.56 Hz) | 1.56 Hz (0.78-1.56 Hz) | p=1.000 | none |
| s18 | 1.56 Hz (0.78-1.56 Hz) | 1.56 Hz (1.56-1.56 Hz) | p=1.000 | none |

**Table S6:** STN burst duration.

| Subject | Off medication (median: IQR) | On medication (median: IQR) | Statistic | Change |
| --- | --- | --- | --- | --- |
| s02 | 353.52 ms (246.09-503.91 ms) | 359.38 ms (246.09-541.02 ms) | p=0.764 | none |
| s04 | 378.91 ms (242.19-532.23 ms) | 425.78 ms (261.72-574.22 ms) | p<0.05 | increase |
| s05 | 339.84 ms (207.03-480.47 ms) | 414.06 ms (253.91-609.38 ms) | p<0.001 | increase |
| s06 | 449.22 ms (261.72-628.91 ms) | 371.09 ms (234.38-519.53 ms) | p<0.001 | decrease |
| s07 | 396.48 ms (257.81-578.12 ms) | 382.81 ms (257.81-563.48 ms) | p=0.593 | none |
| s08 | 421.88 ms (285.16-568.36 ms) | 410.16 ms (257.81-617.19 ms) | p=0.565 | none |
| s10 | 373.05 ms (250.00-509.77 ms) | 394.53 ms (277.34-554.69 ms) | p=0.087 | none |
| s15 | 320.31 ms (199.22-478.52 ms) | 304.69 ms (200.20-444.34 ms) | p=0.285 | none |
| s16 | 408.20 ms (256.84-547.85 ms) | 437.50 ms (281.25-613.28 ms) | p=0.101 | none |
| s17 | 480.47 ms (324.22-675.78 ms) | 378.91 ms (261.72-486.33 ms) | p<0.001 | decrease |
| s18 | 324.22 ms (199.22-457.03 ms) | 330.08 ms (226.56-534.18 ms) | p=0.705 | none |

**Table S7:** STN burst peak amplitude.

| Subject | Off medication (median: IQR) | On medication (median: IQR) | Statistic | Change |
| --- | --- | --- | --- | --- |
| s02 | 0.56 μV (0.40-0.74 μV) | 0.65 μV (0.45-0.96 μV) | p<0.001 | increase |
| s04 | 0.59 μV (0.45-0.82 μV) | 0.82 μV (0.60-1.13 μV) | p<0.001 | increase |
| s05 | 0.54 μV (0.36-0.74 μV) | 0.77 μV (0.54-1.08 μV) | p<0.001 | increase |
| s06 | 1.07 μV (0.69-1.50 μV) | 0.62 μV (0.42-0.86 μV) | p<0.001 | decrease |
| s07 | 0.85 μV (0.62-1.09 μV) | 1.01 μV (0.77-1.45 μV) | p<0.001 | increase |
| s08 | 1.11 μV (0.77-1.62 μV) | 1.06 μV (0.70-1.64 μV) | p=0.366 | none |
| s10 | 0.77 μV (0.53-1.08 μV) | 0.96 μV (0.69-1.37 μV) | p<0.001 | increase |
| s15 | 0.44 μV (0.31-0.65 μV) | 0.39 μV (0.27-0.58 μV) | p=0.011 | decrease |
| s16 | 0.86 μV (0.60-1.16 μV) | 1.10 μV (0.82-1.44 μV) | p<0.001 | increase |
| s17 | 0.84 μV (0.54-1.33 μV) | 0.65 μV (0.46-0.92 μV) | p<0.001 | decrease |
| s18 | 0.44 μV (0.31-0.62 μV) | 0.71 μV (0.46-1.06 μV) | p<0.001 | increase |

**Table S8:** STN burst peak frequency.

| Subject | Off medication (median: IQR) | On medication (median: IQR) | Statistic | Change |
| --- | --- | --- | --- | --- |
| s02 | 17.06 Hz (16.28-20.56 Hz) | 17.44 Hz (15.89-20.17 Hz) | p=0.468 | none |
| s04 | 20.36 Hz (17.44-24.06 Hz) | 21.92 Hz (19.39-24.83 Hz) | p<0.001 | increase |
| s05 | 19.39 Hz (17.06-22.89 Hz) | 17.83 Hz (16.28-20.56 Hz) | p=0.001 | decrease |
| s06 | 20.94 Hz (19.78-22.11 Hz) | 19.78 Hz (17.44-22.50 Hz) | p<0.001 | decrease |
| s07 | 19.39 Hz (17.06-25.22 Hz) | 19.00 Hz (16.28-24.44 Hz) | p=0.610 | none |
| s08 | 20.17 Hz (17.06-22.50 Hz) | 22.50 Hz (19.78-24.83 Hz) | p<0.001 | increase |
| s10 | 17.44 Hz (16.28-20.94 Hz) | 17.44 Hz (15.89-19.78 Hz) | p=1.000 | none |
| s15 | 20.56 Hz (17.06-25.61 Hz) | 20.56 Hz (17.06-24.44 Hz) | p=1.000 | none |
| s16 | 20.94 Hz (17.44-24.44 Hz) | 18.61 Hz (17.06-20.17 Hz) | p<0.001 | decrease |
| s17 | 24.06 Hz (17.83-26.00 Hz) | 19.39 Hz (17.06-23.67 Hz) | p<0.001 | decrease |
| s18 | 18.61 Hz (16.28-22.11 Hz) | 17.44 Hz (15.60-20.17 Hz) | p=0.003 | decrease |

**Table S9:** STN burst frequency span.

| Subject | Off medication (median: IQR) | On medication (median: IQR) | Statistic | Change |
| --- | --- | --- | --- | --- |
| s02 | 1.56 Hz (0.78-1.56 Hz) | 1.56 Hz (1.17-1.56 Hz) | p=1.000 | none |
| s04 | 1.56 Hz (0.78-1.56 Hz) | 1.56 Hz (1.56-2.33 Hz) | p=1.000 | none |
| s05 | 1.56 Hz (0.78-1.56 Hz) | 1.56 Hz (1.56-1.56 Hz) | p=1.000 | none |
| s06 | 1.56 Hz (1.56-1.56 Hz) | 1.56 Hz (0.78-1.56 Hz) | p=1.000 | none |
| s07 | 1.56 Hz (1.56-1.56 Hz) | 1.56 Hz (1.56-1.56 Hz) | p=1.000 | none |
| s08 | 1.56 Hz (1.56-1.56 Hz) | 1.56 Hz (1.56-2.33 Hz) | p=1.000 | none |
| s10 | 1.56 Hz (0.78-1.56 Hz) | 1.56 Hz (1.56-1.56 Hz) | p=1.000 | none |
| s15 | 1.56 Hz (0.78-1.56 Hz) | 1.56 Hz (0.78-1.56 Hz) | p=1.000 | none |
| s16 | 1.56 Hz (1.56-1.56 Hz) | 1.56 Hz (1.56-2.33 Hz) | p=1.000 | none |
| s17 | 1.56 Hz (1.56-1.56 Hz) | 1.56 Hz (0.78-1.56 Hz) | p=1.000 | none |
| s18 | 1.56 Hz (0.78-1.56 Hz) | 1.56 Hz (0.78-1.56 Hz) | p=1.000 | none |

**Figure S1.**
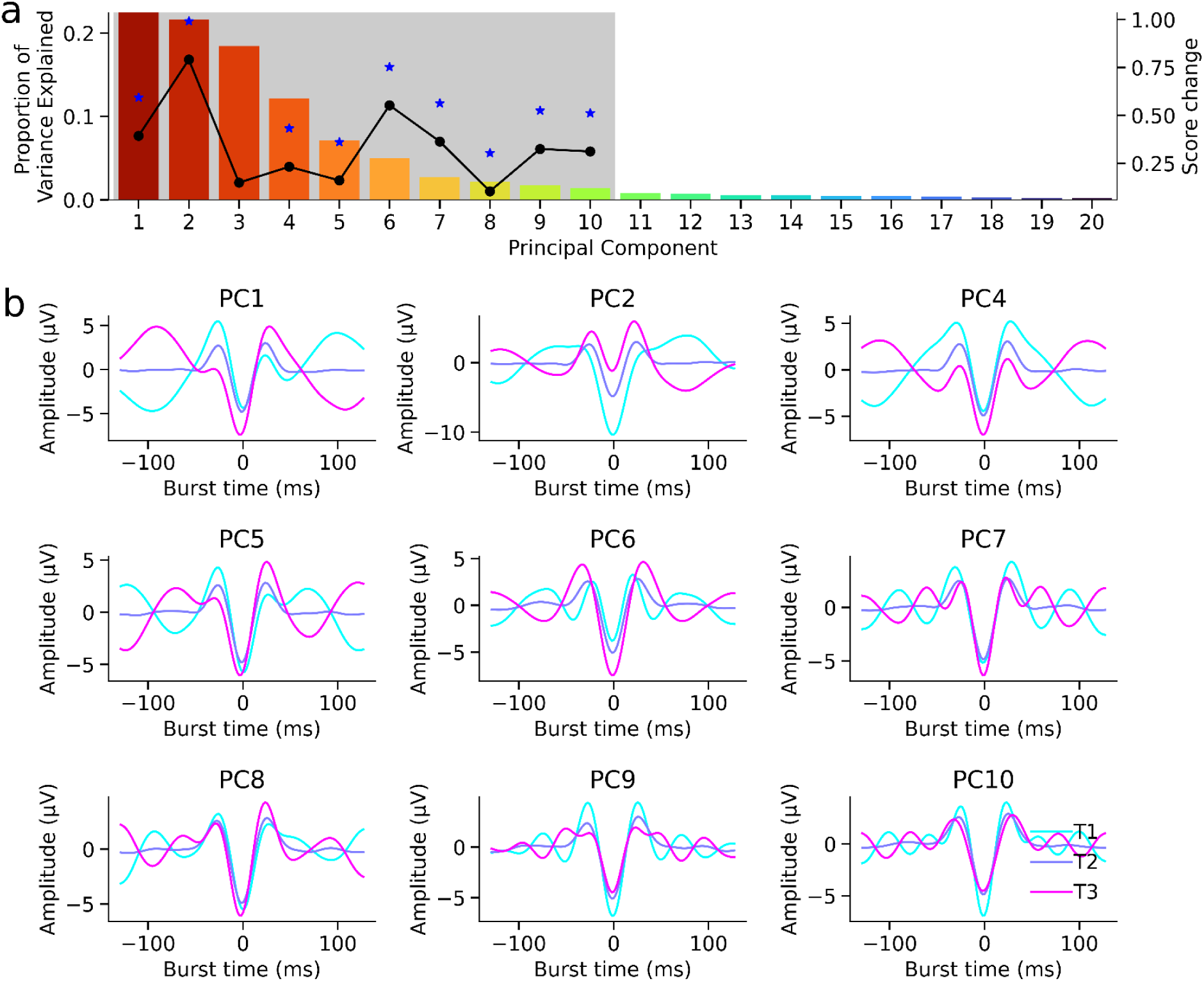
PCA reveals a variety of burst waveform motifs modulated by levodopa. Analysis including all STN contacts. a) The histogram shows the proportion of the variance explained by each component. The grey shaded region shows the significant components identified by the waveform shuffling permutation analysis. The black line overlaid on top of the histogram shows the mean absolute change in score between the OFF- and ON-levodopa conditions. The blue stars indicate the PCs which exhibited significant changes in score. b) For each significant component, mean burst waveform shapes of bursts with a score within the first (cyan), second (blue), and third (magenta) tertile of all burst scores for that component.

**Figure S2.**
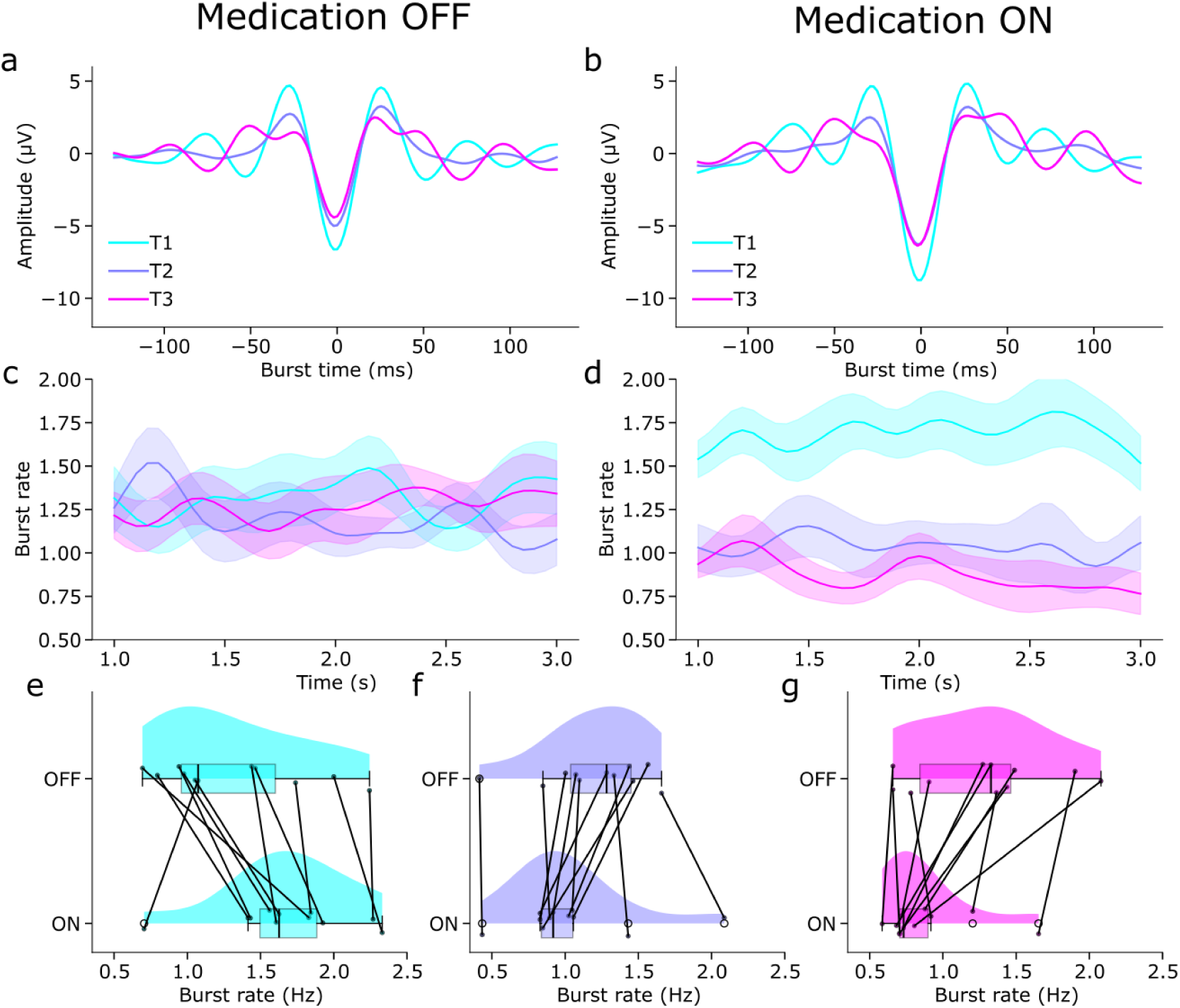
Medication-induced shifts in waveform-specific sensorimotor cortex burst rates along PC9. Analysis including all STN contacts. a) Mean burst waveform shapes of sensorimotor cortex bursts (f during medication off with a score within the first (cyan), second (blue), and third (magenta) tertile of all burst scores for PC9. b) As in a, for bursts during medication on. c) Mean smoothed burst rate (shaded area indicates standard error) across patients off medication over time for each tertile of PC9. d) As in c, during medication on. e-g) Mean burst rate across the entire epoch for the first (e), second (f), and third (g) tertiles of PC9. Points represent individual subjects and black lines connect data points from the same subject.

**Figure S3.**
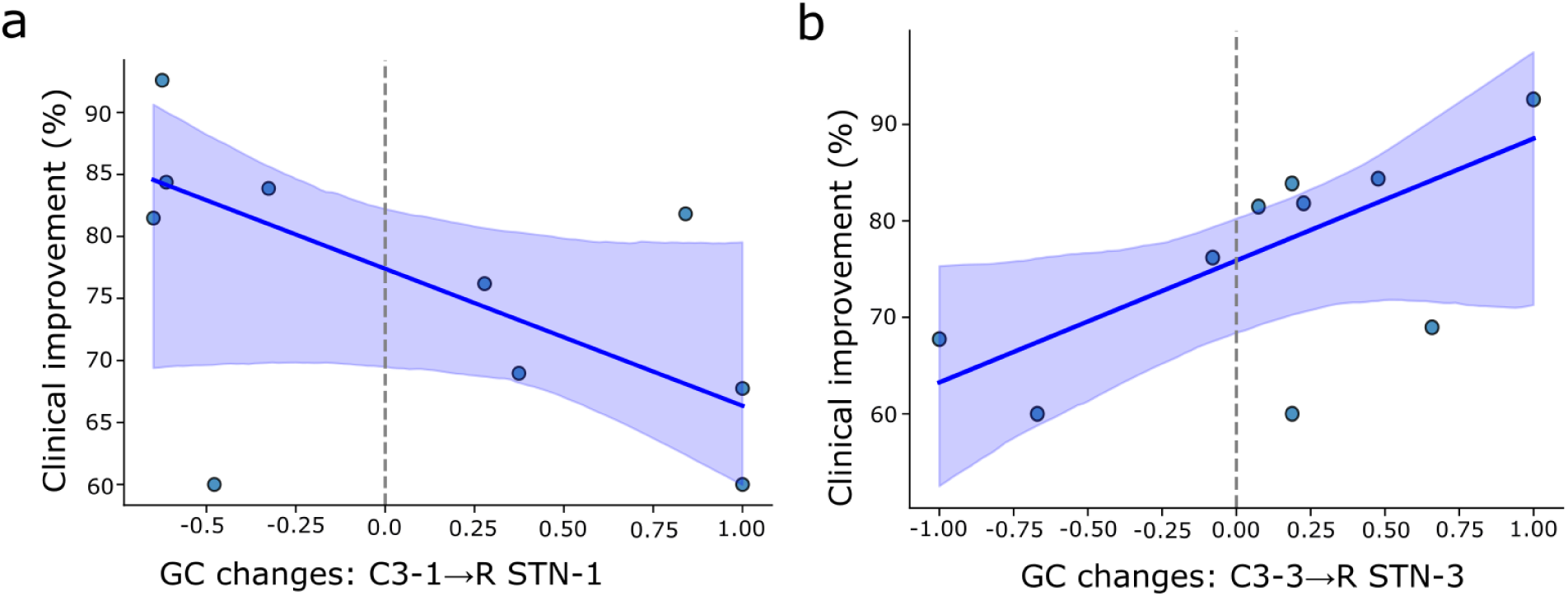
Clinical correlation with waveform-specific FC between the sensorimotor cortex and STN. Bursts were stratified by waveform shape based on tertiles of PC9 loading. a) Changes in FC between the sensorimotor cortex and the STN (best contacts) during first-tertile bursts were negatively correlated with clinical improvement (robust slope =-11.05, *p* = 0.042). b) In contrast, changes in FC between the sensorimotor cortex and the STN during third-tertile bursts were positively correlated with clinical improvement (robust slope = 12.64, *p* = 0.038). Each point reflects one subject; solid lines indicate the fitted linear model, and shaded areas denote 95% confidence intervals.

## Notes

### Competing Interest Statement

The authors have declared no competing interest.

### Funding Statement

This study was funded by Agence Nationale de la Recherche (ANR-18-CE37-0018) SENCE and Ecole Centrale Méditerranée.

### Author Declarations

Ethics committee/IRB of Comité de Protection des Personnes (CPP) Sud Méditerranée I gave ethical approval for this work. The registration number is RCB: 2009-A00913-54.

